# Insights into heterozygous ITPR1 variants associated with ataxia and miosis

**DOI:** 10.1101/2025.04.15.25325838

**Authors:** Josephine Wincent, Songbai Zhang, Andrew Nolan, Frida Nordin, Malin Kvarnung, Per Uhlén, Martin Paucar, Ilse Eidhof

## Abstract

**Background:** Only twice have variants in the ITPR1 gene been described among patients with ataxia and miosis. Functional characterization of these variants is lacking.

**Objective:** To characterize a family affected by congenital ataxia and miosis associated with a novel *ITPR1* variant and to provide a functional assessment for it and two previously reported variants.

**Methods:** Clinical characterization, genetic investigations, and segregation were performed. A novel variant c.7697T>C in *ITPR1* was identified, HEK cells were transfected with vectors carrying our variant and two other previously published variants associated with ataxia and miosis.

**Results:** Ataxia was non-progressive in the reported family, the c.7697T>c ITPR1 variant segregated with disease. Functional validation showed that all the three ITPR1 variants were associated with reduced intracellular calcium release.

**Conclusions:** Here, we present for the first time evidence of pathogenicity for 3 heterozygous *ITPR1* variants in association with ataxia and miosis. Despite being localized in different ITPR1 protein domains, these variants converged on common functional defects.

## Introduction

Spinocerebellar ataxias (SCAs) are a group of clinically heterogeneous hereditary movement disorders with cerebellar ataxia and extra-cerebellar central nervous system manifestations which vary by specific genetic type (Shakkottai & Fogel, 2013). Genetic analyses have led to improved disease classifications, which have enabled the association of SCA with specific genetic disturbances.

*ITPR1* on chromosome 3p26.1 encodes IP3R1, a ligand-gated Ca^2+^ channel (inositol 1,4,5-trisphosphate receptor type 1), localized to the membrane of the endoplasmic reticulum (ER). It is highly expressed in the cerebellum, particularly in Purkinje cells, where it regulates ER-stored Ca^2+^ release in response to the binding of the intracellular second messenger inositol trisphosphate (IP_3_). It is also expressed elsewhere in the brain, including the cerebral cortex, hippocampus, basal ganglia, and thalamus. IP_3_R1, a 2758 amino acid protein, consists of four functional domains including an N-terminal supressor domain and IP_3_-binding domain, a central regulatory domain, and a C-terminal transmembrane/channel domain (Huang et al., 2012; Tada et al., 2016; Tolonen et al., 2024). Pathogenic variants in ITPR1 cause a broad phenotypic spectrum of symptoms depending on the location and effect of the variant(s) on protein function.

Spinocerebellar ataxia type 15 (SCA15 [OMIM #606658]), and spinocerebellar ataxia type 29 (SCA29 [OMIM #117360]) are inherited in an autosomal dominant pattern but the phenotypes are distinguished by age of onset and the presence of cognitive impairment in SCA29. Gillespie’s syndrome (GLSP [OMIM #206700]) may be inherited autosomal dominantly or autosomal recessively and is distinguished clinically from SCA15 and SCA29 by the presence of aniridia (Keehan et al., 2021; Munoz Cardona & Lopez Mahecha, 2022)

ITPR1 variants have previously been associated with ataxia and miosis in two cases (Casey et al., 2017; Chesneau et al., 2024). Here we report the third case, including clinical, genetic and functional characterization using HEK cells and compare it with the previously reported cases.

## Material and methods

### Ethical considerations

This study was approved by Swedish Ethical Review Authority (EPN dnr 2016/2538-32). Informed written and oral consent was obtained from the patients for participation in this study.

### Clinical Investigations

The medical records for each family member were reviewed. Ocular motility was assessed using an eye tracker (VisualEyes 525) and clinical observation. The iris of the index case was examined with slit-lamp and optical coherence tomography (OCT).

### Massive parallel sequencing and Sanger sequencing

Massive parallel sequencing (MPS) of the patient was performed using a 30× PCR-free paired-end WGS protocol on an Illumina NovaSeq 6000 platform as described previously (Magnusson et al., 2020). A gene panel of 956 genes associated with movement disorders and neuromuscular disease was analyzed. The variants were prioritized based on conservation, frequency in internal and public databases, and pattern of inheritance. The ranked variants were then visualized in the Scout analysis platform (Stranneheim et al., 2021). Genes associated with mirror movements were also analyzed (*DCC, RAD51, NTN1*, and *ARHGEF7*). The *ITPR1-*variant was segregated in the family by PCR and Sanger sequencing. The Sanger sequencing was performed by standard methods on an ABI 3730 PRISM® DNA Analyzer. Primer sequences available upon request.

### Plasmids

To construct EGFP-IP_3_R1-WT, the *NheI*-EGFP-IP_3_R1-*XhoI* fragment (restriction enzymes are italicized, same as below) was cut from EGFP-mIP_3_R1-N (Nakayama et al., 2004) and inserted into CAG-MCS2 (Kawauchi et al., 2005). The site-directed mutants of EGFP-IP_3_R1were generated using Pfu Turbo DNA Polymerase (Cat. 600250, Agilent Technologies, Santa Clara, CA, USA) following the manufactures protocol. To generate EGFP-IP_3_R1-R36C and EGFP-IP_3_R1-R36C, the fragment *KpnI*-EGFP-IP_3_R1/N-*NheI* fragment was cut from EGFP-IP_3_R1-WT and inserted into pcDNA-mRFP-GIT1 (Zhang et al., 2009), resulting in the plasmid pcDNA-IP_3_R1/N-EGFP. PCR for IP_3_R1/N-R36C and IP_3_R1/N-R36P was performed using pcDNA-IP3R1/N-EGFP as the template with the following primers (underline indicates mutated nucleic acid, same as below): 5⍰-GGCTTGGTTGATGAC<u>T</u>GTTGTGTTGTACAGC-3⍰, 51-CTTGGTTGATGACC<u>C</u>TTGTGTTGTACAGC-3⍰, respectively. The fragments of *KpnI*-IP_3_R1/N-R36C-EGFP-*NheI* and *KpnI*-IP_3_R1/N-R36P-EGFP-*NheI* were confirmed by sequencing and replaced into EGFP-IP_3_R1-WT, resulting in EGFP-IP_3_R1-R36C and EGFP-IP_3_R1-R36P, respectively. To generate EGFP-IP_3_R1-F2620S, the *EcoRI*-IP_3_R1/C-*XhoI* fragment was cut from GFP-mIP_3_R1-N and inserted into pcDNA-mRFP-GIT1, resulting in the plasmid pcDNA-IP_3_R1/C. PCR for IP_3_R1/C-F2620S was performed using pcDNA-IP_3_R1/C as the template with the following primer: 5⍰-GGCTTGGAAAGGGACAAGT<u>C</u>TGACAATAAGACTGTCACC-3⍰. The *EcoRI*-IP_3_R1/C-F2620S-*XhoI* fragments was confirmed by sequencing and replaced into GFP-mIP_3_R1-N, resulting in GFP-mIP_3_R1-F2620S-N. EGFP-IP_3_R1-F2620S was produced by insertion of the *NheI*-GFP-mIP_3_R1-F2620S-*XhoI* fragment from GFP-mIP_3_R1-F2620S-N into CAG-MCS2.

### Cell culture

HEK-293T cells were purchased (Sigma-Aldrich, Cat. 12022001-1VL) and maintained in DMEM medium (Gibco, Cat. 31966-021) supplemented with 10% fetal bovine serum (FBS, Invitrogen, Cat. 25149-079) and 1% Antibiotic-Antimycotic (Gibco, Cat. 15240062).

### Ca^2+^ imaging

3 ×10^4^ cells per well were seeded in an µ-Plate 96 Well Black ibiTreat (Ibidi, Cat. 89626) coated with Poly-L-Lysine (Sigma, Cat. P4707). One day later, transfection was performed using 57 ng of cDNA per well with 0.28 ml per well of Lipofectamine™ 2000 Transfection Reagent (Invitrogen, Cat. 11668027, same as below) according to the manufacturer’s protocol. After two additional days, the functionality of IP3 R was assessed using Ca^2+^ imaging. In brief, cells were loaded with the Ca^2+^-sensitive fluorescent indicator RHOD4™-AM (10 µM; AAT Bioquest, Cat. ABD-21122) in the presence of 0.04% Pluronic F-127 (ThermoFisher Scientific, Cat. P3000MP) and incubated for 20 min at 37°C in 1x Krebs-Ringer buffer. The buffer was composed of: NaCl (119 mM, Cat. S7653), KCl (2.5 mM, Cat. P5405), NaH_2_PO_4_ monobasic (1 mM, Cat. S3139), CaCl_2_·2H_2_O (2.5 mM, Cat. C3306), MgCl_2_·6H_2_O (1.3 mM, Cat. M2393), HEPES (20 mM, Cat. H4034) and D-Glucose (11 mM, Cat. G8270), with pH adjusted to 7.4 (all from Sigma-Aldrich). Following dye incubation, cells were washed to remove excess dye and incubated for an additional 20 min in 1x KREBS Ringer buffer. Subsequently, the buffer was replaced with 1x Ca^2+^ free Krebs-Ringer buffer containing 2mM EGTA (Sigma-Aldrich, Cat. E4378), in which CaCl_2_·2H_2_O was omitted. Ca^2+^-measurements were performed at 37°C using a Nikon CrEST X-Light V3 inverted confocal spinning disk microscope with a 20x/0.8 dry lens (Nikon). Excitation was assessed at 477 nm and 546 nm for all genotypes (4 × 2 wells) simultaneously, for 15 min at a sampling frequency of 0.5 Hz/well. The equipment was controlled with, and imaging data was collected using NIS Elements software (Nikon). After 5 min, cells were stimulated with either 5 *μ*M ATP (Sigma-Aldrich, Cat. A9187) or 0.5 *μ*M Thapsigargin (ThermoFisher Scientific, Cat. T7459). FIJI, MATLAB (R2021a, MathWorks, USA) and FluoroSNNAP (Patel et al., 2015) were used to process and analyze the collected data.

### Western blot

8 ×10^4^ cells per well were seeded in a 6-well-plate (Falcon, Cat. 353046). One day later, transfection was performed using 1000 ng of cDNA per well with 5 ml per well of Lipofectamine™ 2000 Transfection Reagent. After two additional days, the cells were collected, lysed and sonicated in lysis buffer (10 mM HEPES, pH 7.4, 100 mM NaCl, 2 mM EDTA, 1 mM 2-mercaptoethanol, 0.5% Triton X-100) containing Halt™ Protease and Phosphatase Inhibitor Cocktail (Thermo Scientific, Cat. 78441). The lysates were centrifuged at 20,000 × g at 4 °C for 10 minutes, and the supernatants were collected. Proteins were eluted by boiling in 4 x SDS-PAGE sample loading buffer at 95 °C for 5 minutes, separated by SDS-PAGE, and transferred to a polyvinylidene difluoride membrane (Millipore, Bedford, MA). The membranes were probed with the following primary antibodies: anti-IP_3_R1 antibody (Cell signal technology, Cat. 8568, 1:500), anti-GFP antibody (Abcam, Cat. Ab290, 1:1000), and anti-GAPDH antibody (Cell signal technology, Cat. 5174, 1:1000). The secondary antibodies used were Goat anti-Mouse IgG (Sigma-Aldrich, Cat. A4416, 1:4000), and Goat anti-Rabbit IgG (Sigma-Aldrich, Cat. A6667, 1:4000). Image acquisition and densitometric analysis of the blots were performed with Bio-Rad Image Lab software V4.0.1 (Bio-Rad, Hercules, CA, USA).

### Immunocytochemistry

1 ×10^4^ cells per well were seeded in a 8-well-culture-chamber (Falcon, Cat. 354108). One day later, transfection was performed using 92 ng of cDNA per well with 0.46 ml per well of Lipofectamine™ 2000 Transfection Reagent. After two additional days, the cells were washed once with PBS, treated at 4 °C for 10 minutes with Methanol pre-cold to −20 °C, permeabilized with P-buffer (0.1% Triton X-100 + 0.1% Tween in PBS) at room temperature for 10 minutes, and then blocked with B-buffer (5% skim milk in P-buffer) at room temperature for 1 hour. The cells were subsequently incubated with the primary antibody, anti-KDEL (Santa Cruz Biotechnology, Cat. sc-58774, diluted at 1:250 in B-buffer), followed by the secondary antibody, anti-Mouse IgG (H+L) Alexa Fluor™ 555 (Invitrogen, Cat. A-21422, diluted 1:500 in B-buffer). Coverslips were mounted using Vectashield Antifade Mounting Medium with DAPI (Vector Laboratories, Cat. H-1200-10).

Fluorescence images were acquired using a Olympus FluoView1000 confocal microscope (Olympus, Tokyo, Japan) and analyzed with Olympus FV10-ASW software.

### Statistical analysis

The results were plotted in PRISM (GraphPad Prism 9). Statistical analysis of live Ca^2+^ imaging was done with a non-parametric Kruskal-Wallis test in combination with a Dunns multiple comparisons post-test. P-values lower than 0.05 were considered as significant, with * p < 0.05, ** p < 0.01, *** p < 0.001, **** p < 0.0001.

## Results

### Clinical description

The index case presented with motor difficulties noticed in infancy. Later on, clumsiness, gait difficulties were noted as well as ataxia and miosis. Upon exam the patient displayed mild axial ataxia, broken pursuit, and miosis with slow dark adaptation. The ataxia is non-progressive, and the SARA score was 5.5 points. The patient displayed mirror movements of moderate character, but there were no pyramidal symptoms, areflexia, learning disabilities, dysmorphism, or systemic features. Brain MRI showed mild WMA but no signs of cerebellar atrophy. OCT showed marked thinning of the iris (**Figure 1A**). Multiple relatives of the index case have a similar clinical picture (**Figure 1B**), but only the index case has mirror movements.

**Figure 1:**
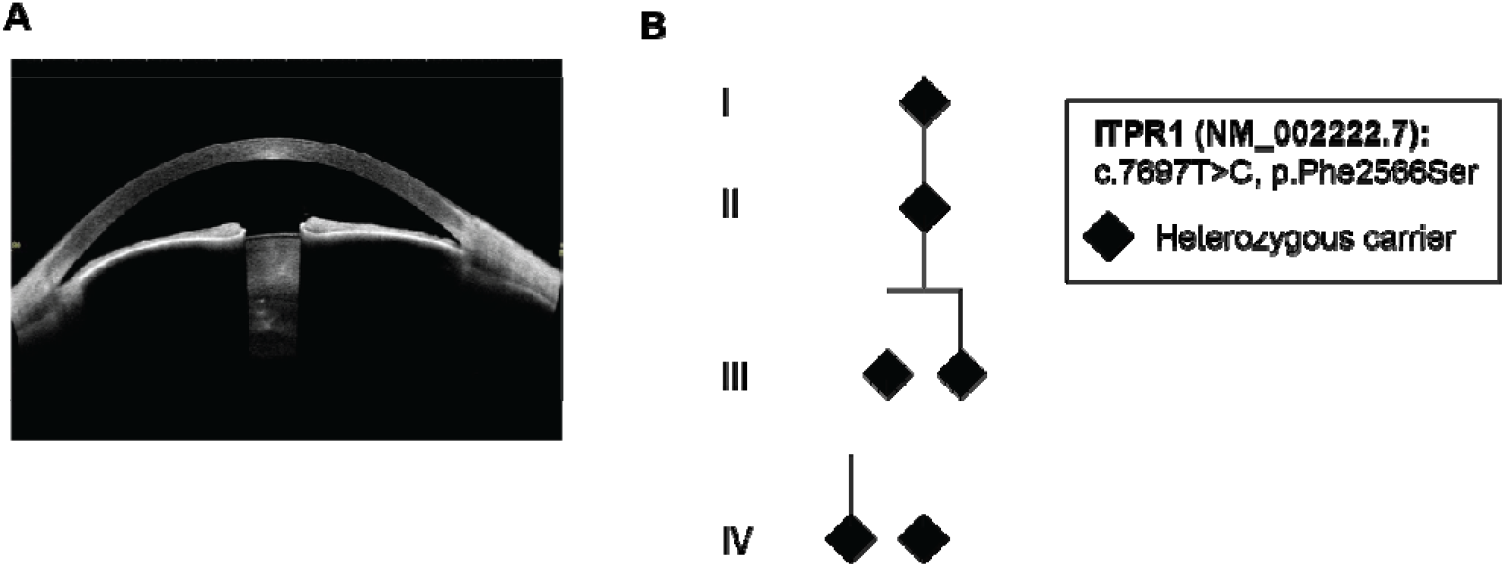
Identification of a novel, heterozygous variant in *ITPR1* in a family with congenital ataxia and miosis. **A**. OCT of the anterior segment of the right eye of index case, revealing a thin iris dilator muscle. **B**. Pedigree showing affected individuals.

### Genetic investigation

The genetic investigation identified a heterozygous missense variant (c.7886T>C, p.Phe2629Ser) in ITPR1 (OMIM 147265, (NM_002222.7)). This novel variant had not previously been reported in the general population (gnomAD v2.1.1) and had a combined annotation-dependent depletion (CADD) score of 32. The affected amino acid was conserved across species according to PHAST, GERP, and phyloP. The missense substitution was predicted to be deleterious by SIFT and probably damaging by Polyphen. The variant is situated in a domain with several previously reported pathological missense variants (observed/expected 0.19 (gnomAD v2.1.1). The variant segregated with the disease in the family (**Figure 1B**) supporting the pathogenicity of the variant. No variants that could explain the patients mirror movement were identified in *DCC, RAD51, NTN1*, or *ARHGEF7*.

### Structural modeling of ataxia-miosis variant in IP_3_R1

The IP_3_R1 is composed of four ITPR1 subunits. Each subunit consists of ten protein domains, including two *β*-trefoil domains (*β*-TF1 and *β*-TF2), three armadillo selenoid folds (ARM1-3), an *α*-helical domain, a intervening lateral domain (ILD), a transmembrane domain (TMD), a helical linker domain (LNK) and a C-terminal domain (CTD) (**Figure 2A**). Ligand binding to IP_3_R1 induces allosteric conformational changes, requiring precise communication between the protein domains. This facilitates channel opening, allowing Ca^2+^ release from the ER into the cytosol. To model the effects of ataxia-miosis variants on IP_3_R1 protein structure, we utilized published cryo-EM rat InP_3_R1 crystal structures (8EAR, 8EAQ and 7LHE). Human and rat IP_3_R1 share 98.44% overall amino acid sequence similarity, including conservation of the residues that are associated with ataxia-miosis variants (*Figure 2A*). These different protein structures enable structural variant analyses under both IP_3_R1 activated (Ca^2+^, IP_3_ and ATP bound: CIA-rIP_3_ R1 (8EAR)) and inhibited (high Ca^2+^ bound: Ca-rIP_3_ R1 (8EAQ), and Ca^2+^ depleted Apo-rIP3R1 (7LHE) configurations.

**Figure 2:**
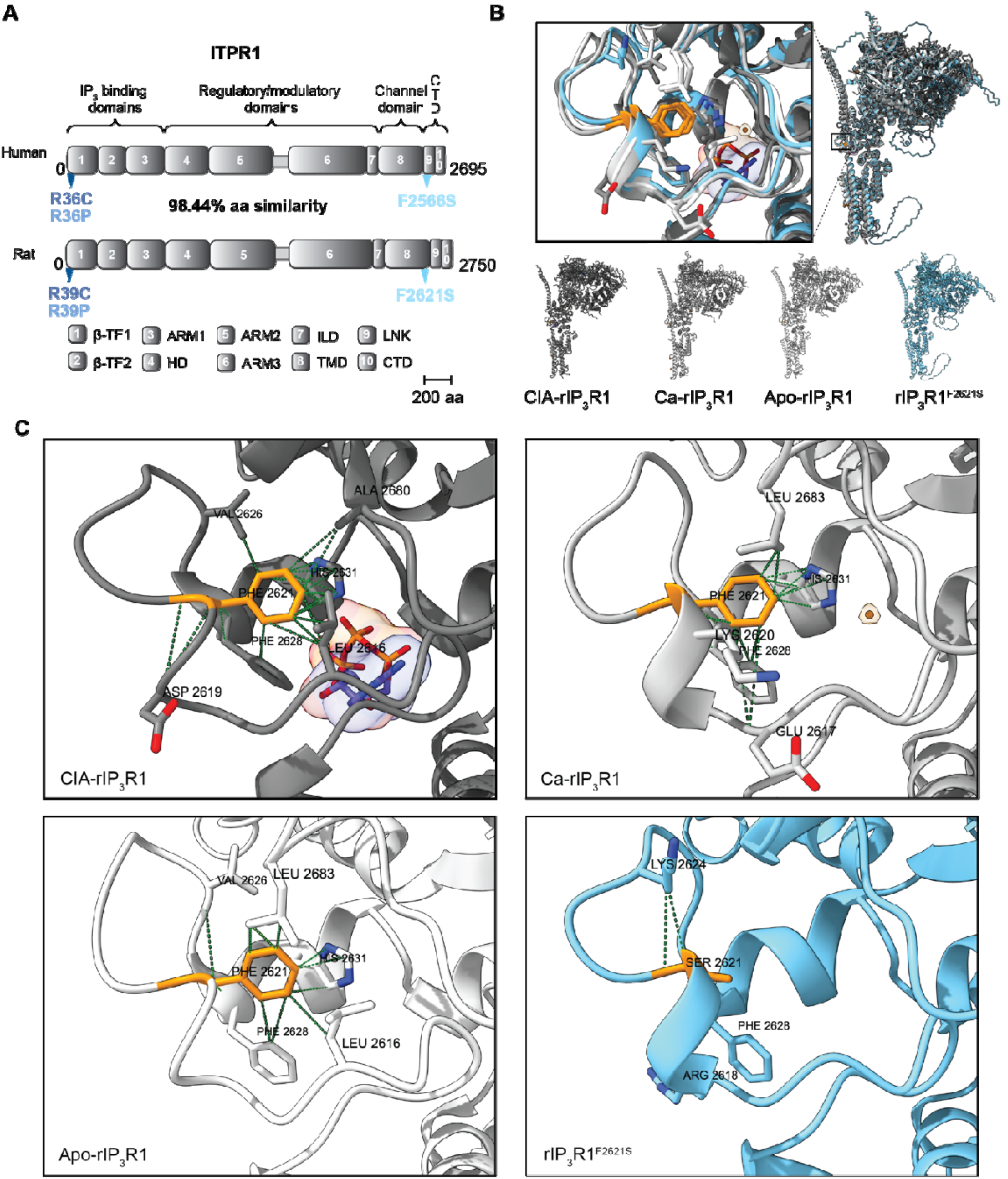
ITPR1 p.Phe2566Ser is located in the ITPR1 CTD domain. **A**. Human ITPR1 and rat ITPR1 are highly similar on protein level. **B**. Chimera overlay of indicated rat IP_3_R1 crystal structures. Zoom in shows Phe2566, corresponding to rat Phe2621 (in orange). **C**. Aminoacid interactions of Phe2621 (in orange) or Ser2621 (in orange) in the indicated structures.

The novel identified p.Phe2629Ser variant is localized in the IP_3_R1 LNK domain, whereas the previously identified ataxia-miosis variants p.Arg36Cys and p.Arg36Pro are located in the *β*-TF1 IP_3_R1 repressor domain (**Figure 2A**). The LNK domain provides a critical link between the CTD and TMD, that together make up the Ca^2+^ gating pore. Upon IP_3_ R1 activation, the LNK domain processes and transmits the ligand-dependent regulatory signals from the cytosolic domains to the Ca^2+^ pore apparatus, suggesting that it is highly relevant for IP_3_ R1 induced Ca^2+^ release.

Under IP_3_R1 ligand-activation conditions, Phe2629, corresponding to Phe2621 in rIP_3_R1, is located in a loop, its side chain pointing towards/facing the inside of the LNK domain (**Figure 2B**). Here it interacts with several different amino acids, such as Leu2616, Val2621, Phe2628, and most predominantly with His2631, which sidechain is at approximately 3.07Å distance. The number of interactions Phe2621 participated in, is decreased in Ca-rIP_3_R1 (around 20) and APO-rIP_3_R1 (around 10) (**Figure 2C**). Suggesting that upon ligand activation, Phe2621 participates in confirmational changes that are required for IP_3_R1 activity.

To investigate whether the p.Phe2621Ser variant could participate in similar types of interactions to any of these IP_3_R1 structures, we used AlphaFold3 to predict the quaternary rIP_3_R1 p.Phe2621Ser protein structure. The overall confidence of the prediction, particularly where the mutation was located, was high. We then mapped the AlphaFold structure (Figure 2B, in blue) onto CIA-rIP_3_R1, Ca-rIP_3_R1 and APO-rIP_3_R1 using Chimera and investigated the location and interactions of Ser2621 (**Figure 2C**). In contrast to CIA-rIP_3_R1 and Ca-rIP_3_R1, Ser2621 was located at the end of an alpha-helix, with its side chain also facing towards the inside of the LNK domain. The number of interactions that Ser2621 participated in, however severely decreased compared to Phe2621 in all IP_3_R1 structures and particularly compared to CIA-rIP_3_R1 (**Figure 2C**). Particularly, the interactions with His2631 seemed to be absent. The only consistent amino-acid interaction was with Phe2628. Moreover, Ser2621, seemed to form new interactions with Arg2618 and Lys2624. We hypothesize that these new interactions and the lack of the original ones, particularly affect the Zn^2+^ binding domain that consists of His2631, His2636, Cys2611 and Cys2614. The exact function of this Zn^2+^ molecule is unknown, but it has been implied in protein folding and providing structural stability. However, interesting to note is that CIA-IP_3_R1 contains a unique ATP-binding pocket located at the interface between the ILD and LNK domain that is absent in both APO-rIP_3_R1 and Ca-rIP_3_R1. Part of the Zn^2+^ finger domain, through C2611 is responsible for the interaction and stabilization of the adenine moiety of ATP in this binding pocket in CIA-rIP_3_R1. ATP binding to this pocket enhances IP_3_ R1 channel activity. The absence of Ser2621 interactions with the Zn^2+^ binding domain, thus likely result in the absence in being able to participate in appropriate ligand-activating and blocking interactions by inducing conformational changes in the Ca^2+^ channel pore.

### Functional analysis of ITPR1 ataxia-miosis variants

We then went on to functionally characterize the novel and previously published ataxia-miosis variants. HEK293T cells were transfected with constructs expressing either wildtype IP_3_ R1, IP_3_ R1^p.R36C^, IP_3_ R1^p.R36P^ or IP_3_ R1^p.F2566S^. These cells still have endogenous levels of IP_3_R1 activity. We confirmed this, and the expression of constructs containing the wild type and mutated proteins (**Figure 3A**). The IP_3_R1 mutations did not seem to affect protein stability or expression (**Figure 3A**). Immunostaining of the GFP-IP_3_R1 mutations expressed in HEK293T cells showed that all IP_3_R1 mutant proteins colocalized with the endoplasmic reticulum marker KDEL, like wild-type IP_3_R1 (**Figure 3B**). This indicates that these mutations do not disturb IP_3_R1 distribution in the ER.

**Figure 3:**
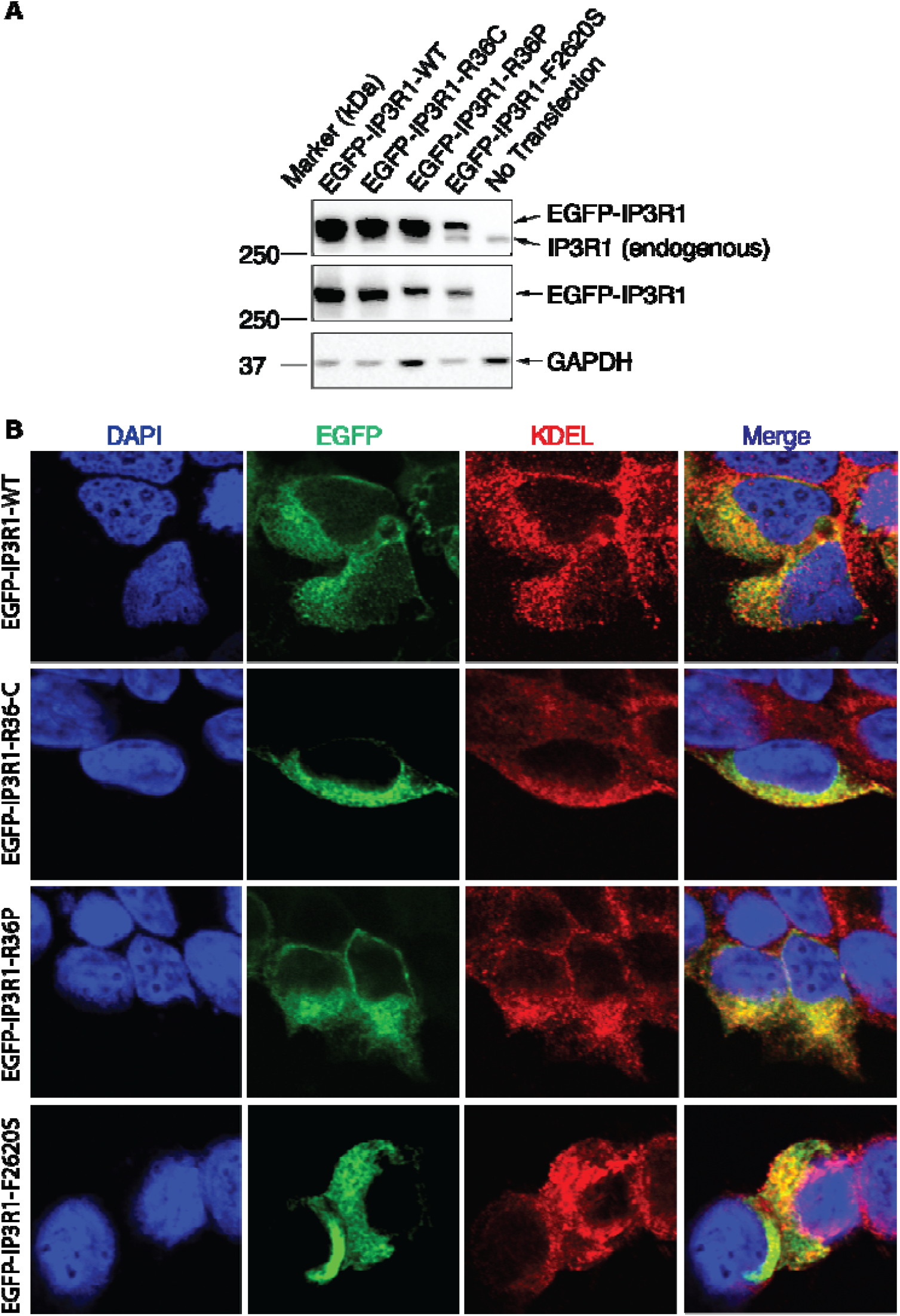
ITPR1 ataxia-miosis variants do not affect IP_3_R1 expression levels nor its localization in the endoplasmic reticulum. **A**. IP_3_R1 western blot of HEK293T cell lines transfected with the indicated constructs. **B**. IP_3_R1-EGFP variants colocalize with the ER marker KDEL in HEK293T cells.

We continued to evaluate IP_3_R1 activity by use of live Ca^2+^ imaging. To examine IP_3_ R1 activity, cells were stimulated with ATP. ATP activates IP_3_R1, through activation of GPCR, leading to cleavage of IP_3_. IP_3_ binding to IP_3_ R1, induces an effect in the receptor, opening the channel pore to let Ca^2+^ flow out of the ER into the cytoplasm (**Figure 4A**). To eliminate the contribution of external Ca^2+^ to this mechanism, these experiments were performed under extracellular Ca^2+^ free conditions. IP3-induced Ca^2+^ release was severely reduced upon all the ataxia-miosis mutations, in a concentration dependent manner (**Figure 4C-D**). Treatment of the cells with Thapsigargin, an inhibitor of the SERCA pump, revealed that intracellular Ca^2+^ levels in the ER were affected upon all mutations (**Figure 4E-G**) Potentially, pointing towards IP_3_R1 channel leakage or a gain-of-function mechanism.

**Figure 4:**
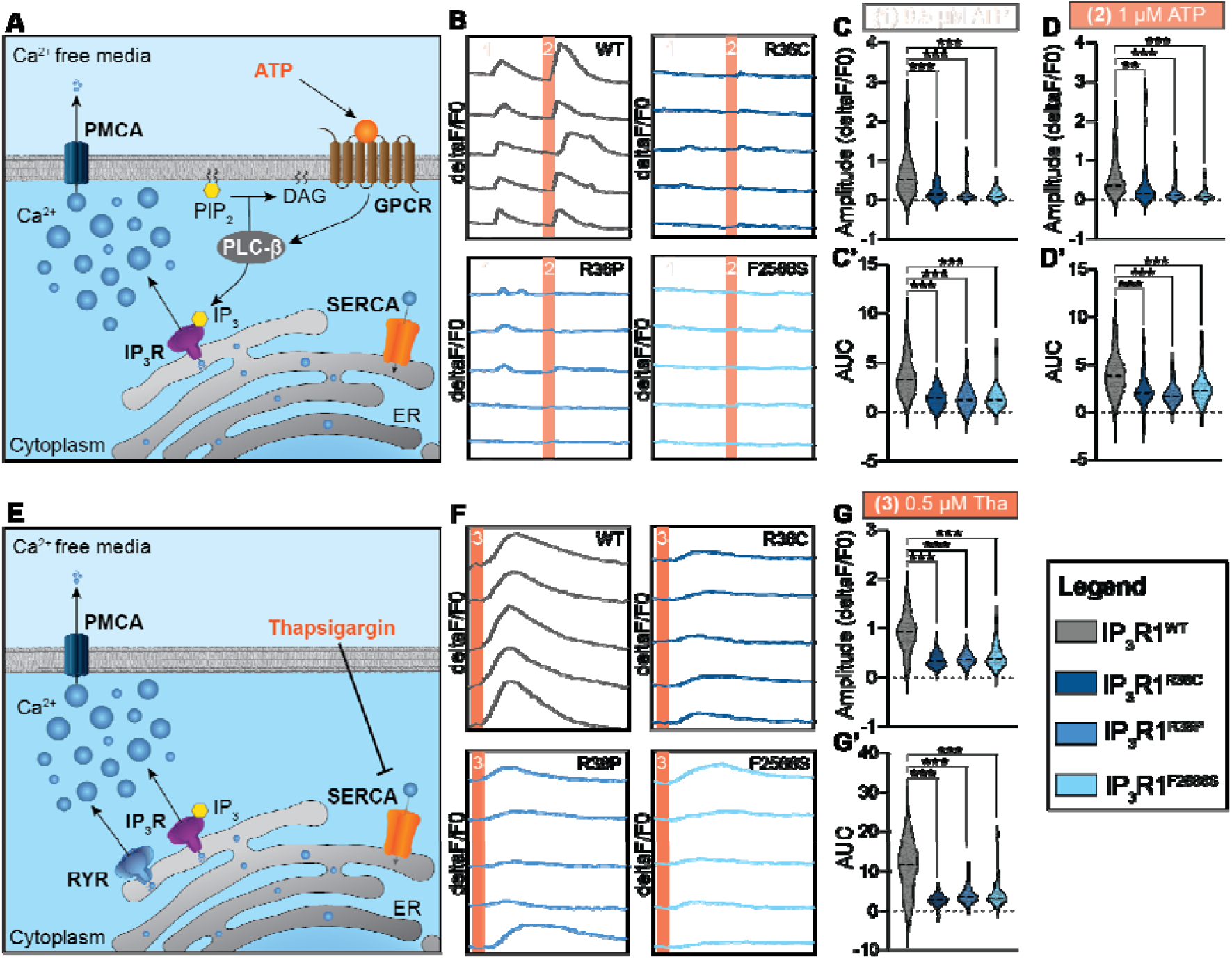
Ataxia-miosis variants affect IP_3_R1 Ca^2+^ release and Ca^2+^ levels in the endoplasmic reticulum. **A**. Scheme displaying IP_3_R1 activation by ATP stimulation. **B**. Representative Ca^2+^ activity trajectories upon 0.5*μ*M ATP (1) and 1 *μ*M ATP (2) stimulation for the indicated genotypes. **C-D**. Quantification of Ca^2+^ peak amplitude (C) and Ca^2+^ levels released from the ER (area-under-Ca^2+^ curve, C’-D’) upon ATP stimulation for the indicated concentrations and genotypes. **E**. Scheme displaying Ca^2+^ release from the ER upon Thapsigargin stimulation. **F**. Representative Ca^2+^ activity trajectories upon 0.5*μ*M Thapsigargin stimulation (3). **G**. Quantification of Ca^2+^ peak amplitude (G) and Ca^2+^ levels released from the ER (area-under-Ca^2+^ curve, G’) upon ATP stimulation for the indicated concentrations and genotypes. **p<0.01, ***p<0.001.

## Discussion

We report here the third case of miosis associated with a pathogenic *ITPR1* variant in a family with early onset ataxia. Miosis has previously been reported twice in association with *ITPR1* variants (Casey et al., 2017; Chesneau et al., 2024). The previously described patients have had a more complex phenotype including mild ID and craniofacial dysmorphism, neither of which was reported in any family member in the current study. However, the iris presentation of our index case is strikingly similar to the iris described in Chesneau et al.’s 2024 study, with a thin dilator muscle without iris transillumination, distinguishing it from congenital microcoria. Interestingly, we found that ITPR1 is strongly expressed in the iris, suggesting that these symptoms could be a direct consequence of ITPR1 function in the eye (**Figure 5A-B**).

**Figure 5:**
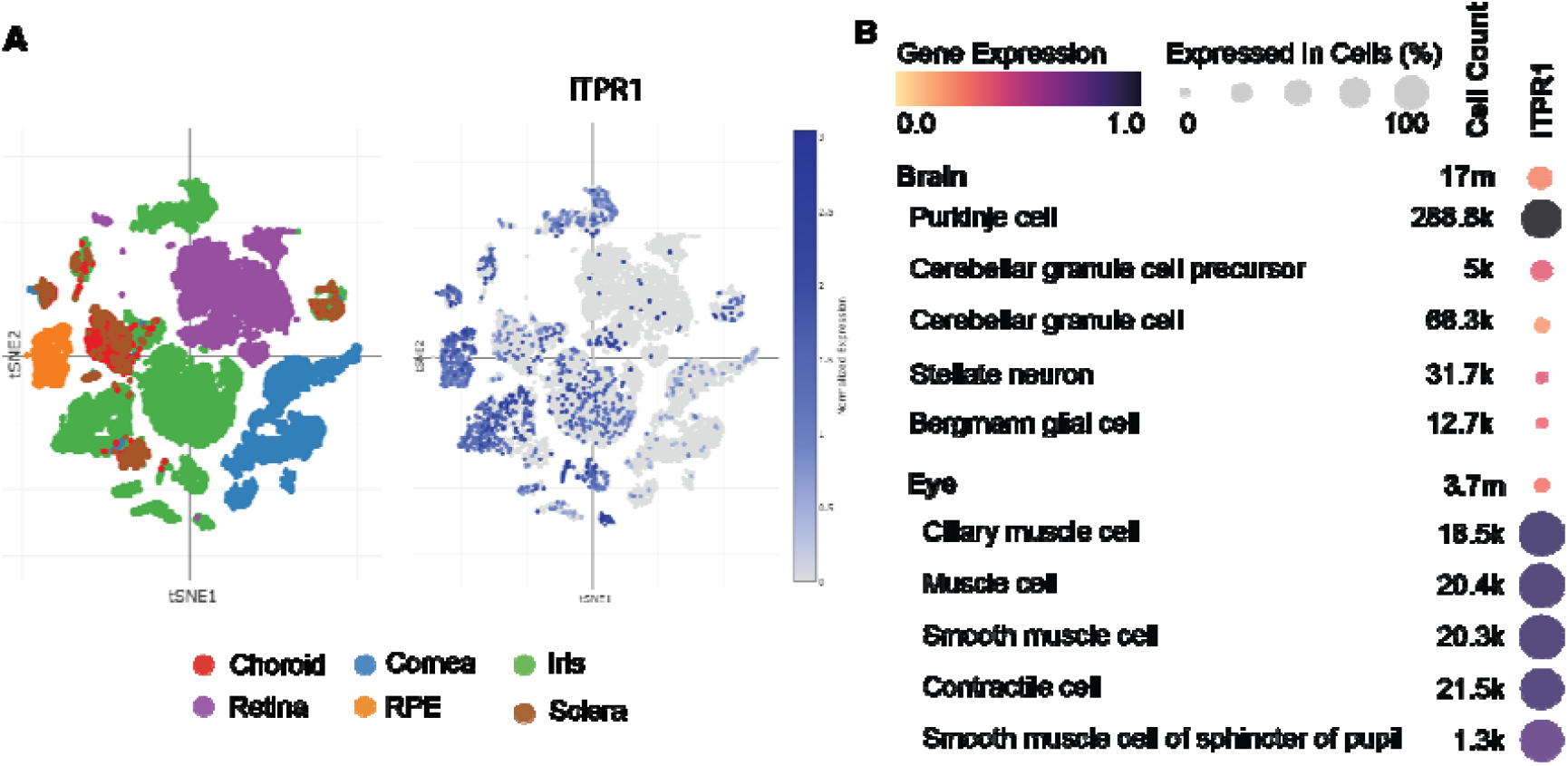
ITPR1 is highly expressed in eye muscle cells and cerebellar Purkinje cells. **A**. tSNE plots showing single-cell ITPR1 expression in the human eye. **B**. Scaled expression of ITPR1 in cerebellar cell types compared to eye cell types that show high ITPR1 expression (from A).

The previously described missense variants were localized in the suppressor domain affecting arginine 36 whereas the current missense variant was localized to the channel domain. Missense variants affecting arginine 36 in individuals with early onset ataxia but no known miosis have also been reported in the medical literature. (Kuperberg et al., 2016; Tolonen et al., 2024) Among the affected family members in the current study one person did not have miosis, also indicating an incomplete penetrance for this ocular sign.

Casey et al. used an IP3 binding assay and single-cell Ca^2+^ imaging to show that the p.Arg36Cys exhibited higher IP_3_-binding affinity and changed the property of the intracellular Ca^2+^ signal from a transient to a sigmoidal pattern, supporting a gain-of-function disease mechanism. Although we did not observe this sigmoidal activity pattern in our functional assays, our findings support that IP_3_R1 variants described in ataxia and miosis may converge on gain-of-function mechanisms.

## Data Availability

All data produced in the present study are available upon reasonable request to the authors.

